## Supplementary material for "Key topics in pandemic health risk communication: A qualitative study of expert opinions and knowledge": S1: S1.docx

**S1 File. Interview guide expert interviews**

**Introduction**

This interview is part of our research on risk communication regarding the coronavirus that causes COVID-19.

In this interview, I am interested in gaining insight into how you think about the prevention, management and risk of coronavirus infection that causes COVID-19.

There are no right or wrong answers, and I don’t have correct answers that I’m looking for. In this interview, your experiences and opinions are important.

I’m going to use a tape recorder, and the audio file will be stored in an encrypted area in my UIS computer that is password protected. Your name will be anonymized when the study is published. What you say will have no consequences for you, and no one other than the researchers involved in the project will have access to the interview data.

**Demography**

- What education/background do you have?
- What is your current position?
- What are your fields of expertise?
- What has been your professional role during the corona pandemic?
- What has been your role in relation to COVID-19 communication? To the public, healthcare professionals, managers?

**Introduction**

1. How do you experience being in a role where you transform knowledge about health risk to the population?
2. What do you think are the main risks associated with the coronavirus that need to be communicated to the public?
3. What are the key concepts that are central for the public to understand when it comes to pandemic risk?

- Early phase
- This phase

**COVID 19- risk and management**

Prompts which apply to question 4-10.

- What considerations have you taken when communicating XX to the public?
- Why are these messages important?
- What considerations have you taken when communicating uncertainty related to XX?
- What do people need to understand related to XX?
- Early in the pandemic versus current phase?

4. What has been important to communicate to the population related to how people are infected by the coronavirus?
5. What has been important to communicate to the public related to the contagiousness of the coronavirus?
6. What has been important to communicate to the public in order to understand the difference in contagiousness and response regarding the seasonal flu and the coronavirus?
7. What has been important to communicate to the public related to how people can limit their exposure to infection?
8. What has been important to communicate to the public in order to avoid being infected and avoid infecting others with the coronavirus?
9. What has been important to communicate to the public when it comes to dealing with the coronavirus?
10. What has been important to communicate to the public regarding the symptoms of COVID-19?
11. What has been important to communicate to the public regarding consequences of COVID-19?

**Jargon**

I have some concepts that have been mentioned in relation to covid communication. I am curious about your opinion and experiences regarding the usefulness of including these terms in public communication.

12. What considerations have been taken in the use of terminology when communicating modes of virus transmission? (contact, droplet, aerosols, surface transmission)?
13. Do you consider the concept exponential growth as a useful concept to communicate risk related to the coronavirus?
    a. What are your experiences with communicating exponential growth?
    b. Why/why not is this concept important?
14. Do you find the R-number useful for communicating risk related to the coronavirus?
    a. What are your experiences with communicating the R-number?
    b. Why/why not is this concept important?

**At the end**

15. Do you have any good advice for our research project? We will make videos that will communicate pandemic risk to the population. What should it be about?
16. In conclusion, is there anything we haven’t talked about related to risk communication during the pandemic that you want to address?
